# Metabolic Dysfunction Related Liver Disease as a Risk Factor for Cancer

**DOI:** 10.1101/2021.10.19.21264255

**Authors:** Alasdair Taylor, Moneeza K Siddiqui, Philip Ambery, Javier Armisen, Benjamin G. Challis, Carolina Haefliger, Ewan R. Pearson, Alex S.F. Doney, John F. Dillon, Colin N.A. Palmer

## Abstract

The aim of this study was to investigate the association with obesity, diabetes and related liver dysfunction and the incidence of cancer.

This study was conducted with health record data available from the National Health Service in Tayside and Fife. GoDARTS, SHARE and Tayside and Fife diabetics, three Scottish cohorts of 13,695, 62,438, and 16,312 patients respectively were analysed in this study. Participants in GoDARTS were a volunteer sample, with half having T2DM. SHARE were a volunteer sample. Tayside and Fife diabetics was a population level cohort. Metabolic dysfunction-related liver disease (MDLD) was defined using ALT measurements, and individuals with alternative causes of liver disease (alcohol abuse, viruses etc) were excluded from the analysis. Other indicators of liver disease were analysed including the Fatty Liver Index, Fibrosis Score(FIB-4) and hospital admissions for NASH. The main outcomes were cancer incidence and cancer death.

MDLD associated with increased cancer incidence with a hazard ratio of 1.31 in a cox proportional hazards model adjusted for sex, type 2 diabetes, BMI, and smoking status (95% CI = 1.27 – 1.35, p < 0.0001). This was replicated in two further cohorts, and similar associations with cancer incidence were found for Fatty Liver Index (FLI), FIB-4 and NASH. Homozygous carriers of the common NAFLD risk variant PNPLA3 rs738409 had increased risk of cancer. (HR = 1.27 (1.02-1.58), p = 3.1×10^−2^). BMI was not independently associated with cancer incidence when MDLD was included as a covariate. MDLD was associated with increased risk of cancer death (HR = 1.40, 95% CI =1.33 - 1.47, p < 0.0001).

MDLD, FLI, FIB-4 and NASH associated with increased risk of cancer incidence and death. Further, we found evidence of a causal association between NAFLD and cancer using the established causal risk allele of PNPLA3 as a genetic instrument. NAFLD may be a major component of the relationship between obesity and cancer incidence.

**Lay Summary:** We found that individuals with metabolic dysfunction-related liver disease (MDLD) have higher overall cancer risk than healthy individuals, as well as increased risk of specific cancers such as colon, breast and lung. We also show that when MDLD is accounted for, obesity does not significantly increase overall cancer risk.

## Introduction

Non-alcoholic fatty liver disease (NAFLD) is the most common cause of liver disease globally, affecting around 25.2% of adults worldwide.^1^ NAFLD, a spectrum of simple steatosis to non-alcoholic steatohepatitis, is traditionally associated with endpoints which affect the liver, including fibrosis, cirrhosis and hepatocellular carcinoma (HCC).^2^ Recent studies have found associations between NAFLD and specific extrahepatic cancers, including colon and breast cancer, as well as overall cancer risk.^3,4^ The relationship between NAFLD and cancer, as well as the synergy between NAFLD and other cancer risk modifiers is not fully understood. Obesity, commonly defined as BMI equal or higher than 30kg/m^2^, is a major cause of NAFLD, with 51.3% of NAFLD patients also being obese.^5,6^ Obesity has also been linked with cancer incidence at 13 different sites in the body.^7^ Wolin et al. estimate that excess weight or obesity accounts for 20% of all cancers.^8^ Mechanistically, several factors associated with increased fat mass have been proposed to cause cancer.^9^ For example, dysregulation of circulating hormones and cytokines including insulin, insulin–like growth factor signalling, adipokines, inflammation and sex hormones may disrupt normal cell cycle control and promote tumour formation. Indeed, there is significant overlap of many of such pathological abnormalities between both overweightness and NAFLD.^10^ The elements of shared pathophysiology of NAFLD and overweightness could potentially mean that the observed increases in cancer risk share a common aetiology. Allen et al. found only small increases in cancer incidence in obese patients without NAFLD. ^4^ Recently a study by Pfister et al. in patients with hepatocellular carcinoma (HCC) showed an associated with NASH and limited anti-tumour surveillance.^11^ This highlights the value in epidemiological studies assessing the relationship between NAFLD and the risk of cancer.

This study utilises an alanine-transaminase (ALT) defined metabolic dysfunction-related liver disease phenotype (MDLD).^12–14^ ALT is positively correlated with hepatic steatosis, and when other causes of liver disease are excluded, this can be an effective method of NAFLD diagnosis. ^15^ Recently Eslam et al. described a phenotype of metabolic dysfunction associated fatty liver disease (MAFLD) based on biochemical, imaging or biopsy along with T2DM, obesity or other metabolic risk factors.^16,17^ The diagnosis of NAFLD in clinical settings often differs from the definitions applied in large scale epidemiological studies. Clinically patients with suspected NAFLD are not always subjected to invasive investigations such as biopsies or ultrasound imaging, therefore this necessitates the use of more commonly measured biomarkers and use of exclusion criteria to eliminate other causes of raised ALT. Therefore in this study we develop and validate a definition of metabolic dysfunction-related liver disease (MDLD) in large scale data resources and in order to test its associations with cancer incidence.^18^

The aim of this study was to analyse the effects of MDLD on cancer incidence and cancer death. In addition, the study aimed to investigate the interaction between BMI and MDLD. Finally, using Mendelian randomisation methods, we investigated whether the relationship between MDLD and cancer was causal or not.

## Methods

### Data

#### GoDARTS (Genetics of Diabetes Audit and Research Tayside, Scotland)

This study aimed to analyse the incidence of all cancer longitudinally. The first cohort used was GoDARTS, a case-control type 2 diabetes study based in Tayside, Scotland. Key descriptive statistics and demographic attributes of this cohort are shown in Table 1. This cohort was used for discovery and comprised of electronic health records (EHRs) from 13,695 eligible individuals.^19^ The mean age at sign up was 63.41 years and participants had a mean follow up of 8.95 years. 48.6% of patients were male. On patients’ date of sign up they were phenotyped by biochemical and haematological investigations, anthropometric measurements and lifestyle questionnaires. This date was used as the beginning of the follow up period. 2,794 patients had cancer incidents during the follow-up period.

**Table 1:** Mean Characteristics of GoDARTS Patients Stratified by NAFLD Status at Time of enrolment to GoDARTS.

| Characteristic | Non NAFLD | Number NAFLD | p |
| --- | --- | --- | --- |
| Number | 6726 | 6969 |  |
| % Diabetic | 24.65%(n = 1,658) | 66.34%(n = 4,623) | < 0.0001 |
| BMI | 27.22kg/m<sup>2</sup>(SD = 4.76) | 30.90kg/m<sup>2</sup>(SD = 6.07) | < 0.0001 |
| Weight (Males/Females) | 84.44kg(SD = 14.18)<br>/70.20kg (SD = 14.38) | 92.89kg(SD = 17.04)<br>/79.64kg(SD = 18.30) | < 0.0001<br>/< 0.0001 |
| Female | 41.48% (n = 2790) | 55.46% (n = 3865) | < 0.0001 |
| Smoker | 55.14% (n = 3709) | 57.67% (n = 4019) | 2.3x10<sup>-3</sup> |
| Age at Signup | 61.88 years(SD = 13.72) | 64.9 years(SD = 11.54) | < 0.0001 |
| Follow-up Length | 9.24years(SD = 2.50) | 8.58 years(SD = 2.88) | < 0.0001 |

#### SHARE (Scottish Health Research Register)

Two further, independent cohorts were analysed for replication. The second data source was SHARE. This is a cohort in which individuals volunteer to allow their medical records to be used for scientific research, and is open to anyone in Scotland over the age of 16. The characteristics of this cohort are shown in table 2.This comprised 62,438 patients with EHRs available once patients with exclusions for alternate causes of raised ALT livers were removed.^20^ This cohort was used for replication of findings in GoDARTS. The mean age in SHARE was 57.0 years, and 61.6% were female.

**Table 2:**
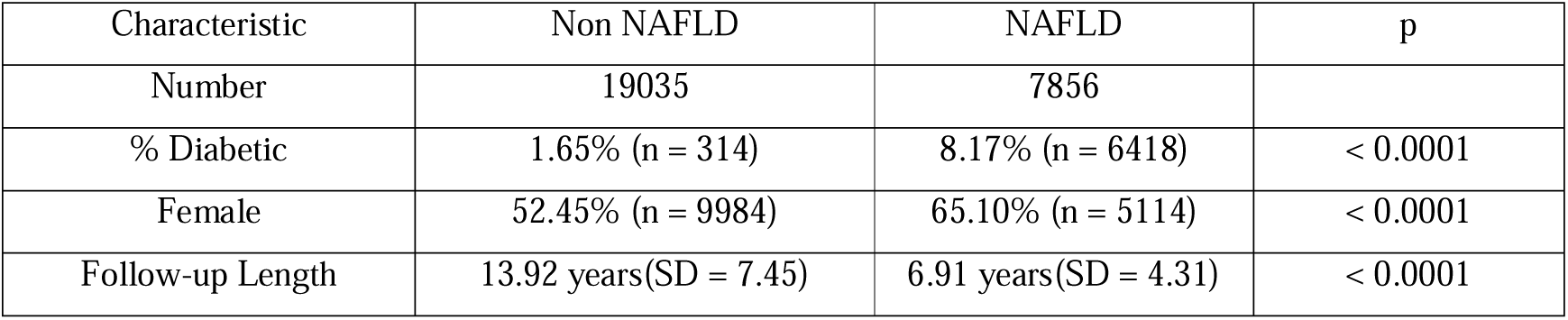
Mean Characteristics of SHARE Patients Stratified by NAFLD Status at Age 60 (Beginning of Follow-Up Period)

| Characteristic | Non NAFLD | NAFLD | p |
| --- | --- | --- | --- |
| Number | 19035 | 7856 |  |
| % Diabetic | 1.65% (n = 314) | 8.17% (n = 6418) | < 0.0001 |
| Female | 52.45% (n = 9984) | 65.10% (n = 5114) | < 0.0001 |
| Follow-up Length | 13.92 years(SD = 7.45) | 6.91 years(SD = 4.31) | < 0.0001 |

#### Tayside and Fife T2DM Cohort

Replication of results was also undertaken in Tayside and Fife T2DM Cohort (T&F). This cohort comprises all patients in the Tayside and Fife NHS region who had a diagnosis of T2DM at some point in their lives. Many of the patients received a diagnosis of T2DM during the follow up period, therefore the T2DM rate is not 100% at baseline. The characteristics of this cohort are shown in table 3. Like the two previous cohorts, medical records from the NHS are available for these patients. The cohort 16,312 patients eligible after exclusions for other hepatic insults were made. The mean age of these patients was 65.0 years, and 48.1% were female. Results for analysis in T&F are reported in the supplemental appendix.

**Table 3:** Mean Characteristics of Tayside and Fife Diabetics Patients Stratified by NAFLD Status at Age 60 (Beginning of Follow-Up Period)

| Characteristic | Non NAFLD | NAFLD | p |
| --- | --- | --- | --- |
| Number | 5,102 | 6039 |  |
| % Diabetic | 40.53% (n = 2068) | 60.37% (n = 3646) | < 0.0001 |
| BMI | 30.55kg/m<sup>2</sup> (SD = 6.12) | 33.15kg/m<sup>2</sup> (SD = 6.78) | <0.0001 |
| Female | 45.84% (n = 2339) | 53.27% (n = 3217) | < 0.0001 |
| Smoker | 58.57% (n = 2988) | 65.01% (n = 3926) | <0.0001 |
| Follow-up Length | 11.08 years (SD = 6.07) | 6.21 years (SD = 4.03) | < 0.0001 |

The results from T&F were not meta analysed with GoDARTS and SHARE, as this is a primarily diabetic cohort, therefore does not capture those who do not go on to get diabetes. The ascertainment bias in this cohort that only contains individuals who did eventually get diabetes is likely to have resulted in the lower point estimate for MDLD in cancer risk that we have observed. To ensure there was no overlap in participants between cohorts, patients in SHARE who were also in GoDARTS were excluded from SHARE, and participants in GoDARTS or SHARE were excluded from analysis in T&F, meaning each cohort was completely independent.

To allow comparison with the GoDARTS cohort, a baseline point had to be chosen from which to begin the follow up period in which to analyse cancer incidence in SHARE and T&F. The age of 60 was chosen as it is close to the mean baseline age of GoDARTS, and importantly close to the mean age of MDLD diagnosis in GoDARTS (60.8 years) and in the literature.^21^ This allowed a more robust replication of findings in GoDARTS in the two replication cohorts, ensuring age wasn’t a source of heterogeneity in analysis. These criteria left 26,891 patients in SHARE and 11,141 patients in T&F suitable for analysis with a median follow up time of 11.0 years and 8.0 years respectively. The EMRs available for patients in all cohorts are from the NHS Tayside and Fife authorities. Consort diagrams can be found for each cohort in the supplemental appendix.

### Outcomes

All outcomes were defined using NHS medical record data, made available for participants in each of the three cohorts. As such, all data was recorded in the same format; all disease was recorded in ICD10 codes and biochemical measures in the same, relevant units.^22^ (E.g. Units per litre for ALT measurements)

### MDLD Phenotype

MDLD cases and controls were defined using the liver function test alanine transaminase (ALT), a commonly used marker of liver damage, and a useful surrogate for NAFLD.^12,14,23–26^ This was chosen as it is commonly measured and Schindhelm et al. found in a population cohort that raised ALTs are a good surrogate for NAFLD.^23^ Elevated ALT levels were considered to be over 30U/L for men, and over 19U/L for women (Normal ALT reference ranges: Males - 5-30U/L; Females – 5-19U/L.). These upper limits are those suggested by Prati et al. as the maximum normal values of ALT in healthy adult men and women.^27^ Raised ALT levels correlate with NAFLD and are an appropriate surrogate marker for the disease, provided other causes of liver disease are ruled out.^23,28^ There is substantial evidence that raised ALT levels in the absence of any apparent liver insult are extremely likely to be caused by NAFLD.^29^ All samples from GoDARTS and SHARE were analysed in the same laboratory.

MDLD cases were defined as any patient who had experienced at least 2 raised ALT measurements, at least 3 months apart. This time scale was chosen as 3 months is a commonly used definition of chronic and most cases of acute hepatitis, such as drug induced, will have resolved.^30^ This also increases the specificity of the definition. Due to the lack of confirmatory biopsy and ultrasound data in all patients, we have refrained from referring to this phenotype as “NAFLD”, and instead have used MDLD.

#### Exclusions

Patients were excluded from analysis if they had features of other chronic liver disease recorded in their medical records. These included: any positive serological tests for anti-smooth muscle antibody, antinuclear antibodies or anti-mitochondrial antibodies, any positive serology for hepatitis B surface antigen or hepatitis C antibody, or mention of cause of liver disease in medical records. In GoDARTS, 1,157 patients had both immunological and virological screens at some point which were negative, therefore they were included in analysis. Patients with alcohol dependence or any documentation of alcoholic liver disease in their EHRs were excluded using ICD codes: “K70” and “F10”. In addition, patients who self-reported drinking more than 20g (2.5 units) a day for women and more than 30g (3.75 units) a day for men were excluded. Allen et al. concluded that alcohol was not likely to explain the increase in cancer incidence seen with NAFLD in their study.^4^

#### Validation of Phenotype

To validate this phenotype, sensitivity and specificity analyses were conducted in GoDARTS comparing this to cases of NAFLD confirmed in EHRs with the “K76.0” ICD10 code. The sensitivity of this definition was 97.4%, and the specificity was 32.0%. These analyses were also conducted in SHARE using the same method, with a sensitivity of 75.3% and specificity of 54.2%, and in T&F with sensitivity of 94.6% and specificity of 38.3%. The SHARE cohort has lower sensitivity compared to the other two cohorts, likely due to the lower average age of the cohort and the lower prevalence of diabetes resulting in lower healthcare interaction, morbidity and mortality. Also, due to the relatively low numbers of confirmed NAFLD in EHRs, small differences in numbers can have large effects on sensitivity and specificity percentages.

Another method of detecting NAFLD non-invasively is the Fatty Liver Index (FLI).^31^ This uses BMI, waist circumference, triglycerides and gamma-glutamyl transferase (GGT) to define NAFLD, and has been validated in a number of cohorts as an accurate surrogate of NAFLD. 4,164 patients in GoDARTS had the required data available for this measure. In GoDARTS, FLI correlated significantly with MDLD as diagnosed by ALT levels. (Pearson correlation coefficient = 0.33 (0.31-0.36), p < 0.0001)

The FIB-4 scoring system was also used in the GoDARTS study.^32^ A FIB-4 score of greater than 3.25 has been shown to predict advanced hepatic fibrosis, therefore this score was used as the cut off. This was calculated using the highest recorded AST and ALT measurements and platelet count before the beginning of the GoDARTS for each individual to calculate the highest FIB-4 score they had experienced.

To further validate this phenotype positive control tests were run against chronic kidney disease (CKD) in GoDARTS, as it has been shown to associate with NAFLD.^33^ During the follow up, 1,131 patients had incidence of CKD. MDLD was found to associate with incidence of CKD in a cox proportional hazards model adjusted for sex, T2DM, age, and BMI. (HR = 1.32(1.25 – 1.39), p < 0.0001)

A positive control test with the well-known NAFLD risk variant *PNPLA3* rs738409 was conducted. ^34^ In GoDARTS, 8,399 eligible participants had been genotyped for this variant. In a logistic regression with an additive model, adjusted for age and sex, *PNPLA3* rs738409 was associated with increased MDLD at the beginning of the study. (OR = 1.23(1.12-1.36), p < 0.0001)

### Clinically Adjudicated NAFLD and NASH

As well as our ALT based MDLD definition, some patients had NAFLD confirmed in hospital admissions data with the ICD10 code “K76.0”. This is referred to as “NAFLD hospitalisation” in subsequent sections. In GoDARTS, 0.36% of participants had this code reported in their medical records at any point.

Non-Alcoholic Steatohepatitis (NASH) was phenotyped by searching admissions, deaths and biopsy files for cases of NASH, defined using the ICD10 codes for NASH, fibrosis and cirrhosis. This may have been a main cause of hospitalisation or concomitant morbidity.

### Mendelian Randomisation

Mendelian randomisation methods were used to assess whether the relationship between MDLD and cancer incidence was causative.^35^ The missense variant *PNPLA3* rs734809, which is strongly associated with the development and progression of fatty liver disease, was chosen as it has been shown in a large number of studies to associate with MDLD, and has been used in previous Mendelian randomisation studies on MDLD. The ratio method was used to conduct this analysis..^34^ In GoDARTS, 7,715 patients had been genotyped for this variant, and 343 of these were homozygous carriers. (Minor Allele Frequency (MAF) = 20.6%) In SHARE, 1,755 patients had been genotyped for this variant, with 50 being homozygous carriers. (MAF = 23.0%)

### Overweight and Obesity Definitions

In this study, overweight is defined as a BMI greater than 25kg/m^2^ and less than 30kg/m^2^. Obesity is defined as a BMI equal or over 30kg/m^2^.^36^

### Cancer Phenotype

Cancer incident data was obtained from the Scottish cancer register, part of the Scottish Morbidity Record.^37^ This contains all diagnoses of cancer made in Scotland in NHS care, in ICD10 code format. This data was available for patients in GoDARTS, SHARE and T&F. Cases were cross checked with recorded cases in hospital admissions and death records files. The cancer records were identified by the relevant ICD10 codes for malignant neoplasms or neoplasms of unknown behaviour. These were any code including “C”, “D0”, “D37”, “D38”, “D39”, or “D4”. Obesity related cancer incidents were phenotyped similarly, but specifically for the 13 reported obesity related cancer sites. ^7^

Cancer deaths were phenotyped based on death certificate files in EHRs. These list a main cause of death and contributing causes of death for each patient who has died. These were also cross checked with the Scottish cancer register file.

### Statistical Methods

All data analysis was carried out in the statistical package R. The effects of MDLD and other independent variables on cancer incidence were analysed using a Cox proportional hazards model (CPH). Patients were censored at the point at which they had a cancer incident recorded, death, or September 2016 when the follow-up period ended. Patients with missing data were excluded from analysis.

To assess whether MDLD affected cancer death risk in the presence of non-cancer death as a competing risk regressions (CRR) using Fine and Gray’s method were run. Logistic regression (LR) models were used to evaluate the effect of MDLD on death cause.

In the GoDARTS cohort models were adjusted for sex, age, BMI, T2DM, and smoking status. In GoDARTs, models with BMI replaced by weight or waist measurement were also run, as these are slightly different measure of obesity and may have provided further insight into the associations. Hypertension, activity level, alcohol consumption and deprivation level were not included in the models as they did not have a significant effect on cancer incidence in the adjusted model. In the SHARE cohort, models were adjusted for sex and T2DM. Smoking and BMI data were not widely available for individuals in the SHARE cohort, therefore this was not controlled for in most analyses.

### Patient and Public Involvement statement

There was no patient involvement in the design of this study.

## Results

### MDLD and Cancer Incidence

In the GoDARTS cohort, MDLD was associated with increased cancer incidence. During the follow up period, 18.5% of controls compared to 22.2% of patients with MDLD developed cancer. In controls, 1244 patients had cancer incidents and 1550 patients had incidents in MDLD cases. Patients who had MDLD at enrolment to GoDARTS had increased cancer incidence independent of sex, age, BMI, smoking status, and diabetes status (Figure 1, HR = 1.31(1.27 – 1.35), p < 0.0001). Using the same covariates, the Fatty Liver Index was associated with increased cancer incidence (HR = 1.004(1.00-1.008), p = 5.0×10^−2^) and FIB-4 score over 3.25 was associated with increased cancer risk. (HR= 1.31, 95% CI =1.29 – 1.53, p = 3.2×10^−3^).

**Figure 1:**
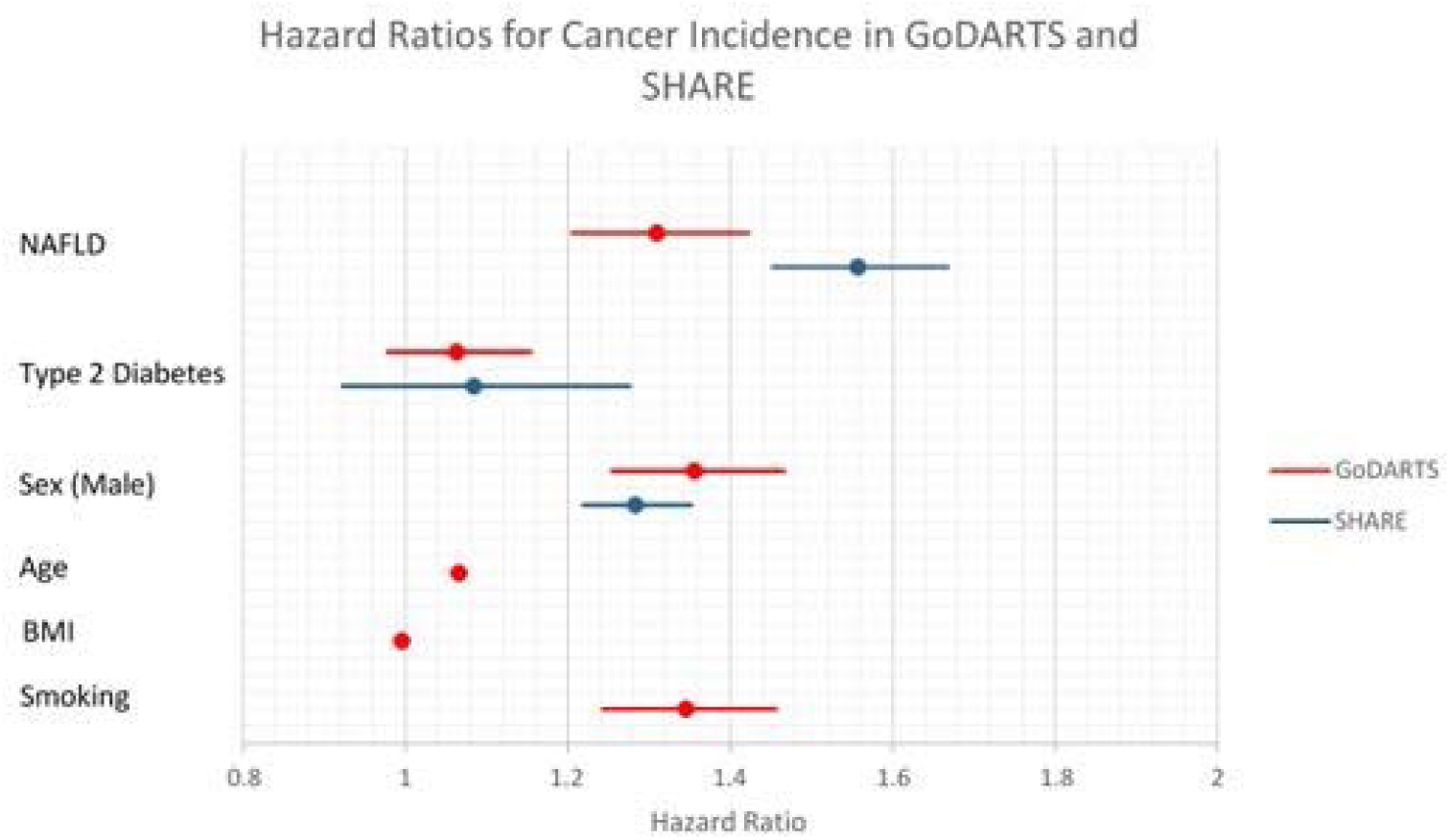
Hazard Ratios for Cancer Incidence in GoDARTS and SHARE.

When MDLD was not taken into account, BMI was associated with increased cancer incidence (HR = 1.09(1.01 – 1.18), p = 3.1×10^−2^). This association was completely abrogated when adjusted for the presence of MDLD. Similar results were found for other markers of adiposity, weight and waist measurements.

When analysis was limited to obesity related cancers, BMI was associated with increased cancer incidence. (HR = 1.01(1.00 – 1.03), p = 3.3×10^−2^) Similarly to the analysis of all cancer incidence, BMI was not associated with cancer incidence when MDLD was added as a covariate.

Similar results were found in the SHARE cohort. Out of 26,891 patients analysed, 5,728 had cancer incidents in the follow up period. MDLD was associated with increased cancer incidence (Figure 1, HR = 1.56, (1.45-1.67), p < 0.0001). MDLD hospitalisation prior to baseline was associated with increased cancer risk, with a hazard ratio of 2.54. (95% CI = 1.14 – 5.65, p = 2.3×10^−2^). NASH was also associated with increased cancer incidence. (HR = 4.18(1.74-10.0), p = 1.4×10^−3^) Among the patients in SHARE, 1,912 had BMI data available. In these patients, when MDLD was accounted for, BMI was not significantly associated with overall cancer incidence, or with obesity related cancer incidence.

Similar results were found in the population based diabetes cohort from Tayside & Fife. Out of the 11,141 patients analysed, 1,819 had cancer incidents in the follow up period after the age of 60. MDLD was associated with cancer incidence in the follow up period. (HR = 1.16(1.04-1.29), p =5.9×10^−3^) Full results are shown in the supplementary appendix.

As well as increasing all primary cancer incidence, MDLD was associated with increased incidence of specific cancers in GoDARTS and SHARE, shown in Figure 2. Due to lower numbers of cases, the confidence intervals for these are wider than for all primary cancers combined. Breast and uterine cancer analyses were limited to females, with prostate cancer analyses limited to males. T&F was not meta analysed in this analysis due to the primarily diabetic composition of the cohort, which did not capture those over 60 who did not go on to get T2DM. The ascertainment bias in this cohort that only contains individuals who did eventually get diabetes is likely to have resulted in the lower point estimate for MDLD in cancer risk that we have observed.

**Figure 2:**
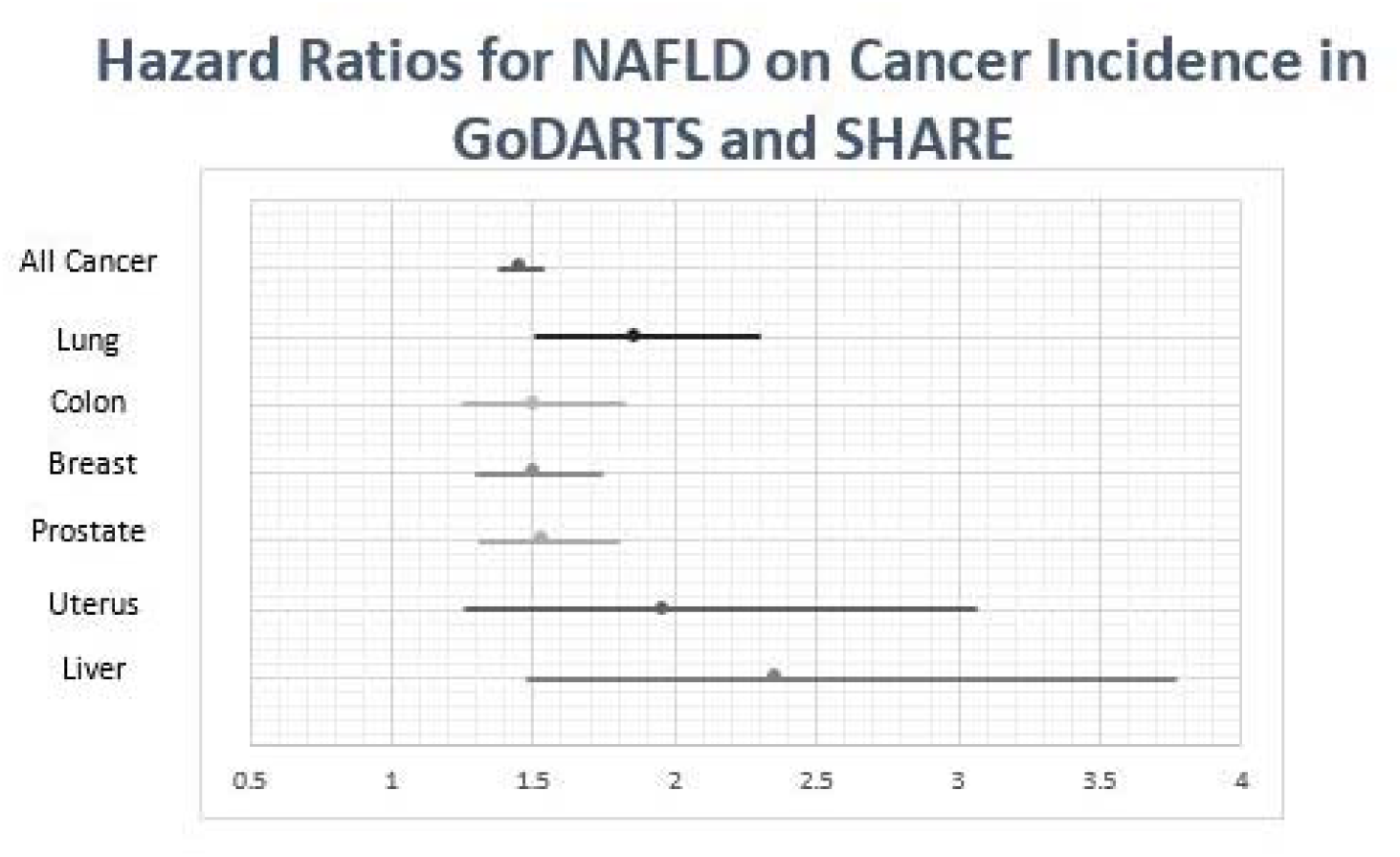
Hazard Ratios for Cancer Incidence at Specific Sites in Meta-Analyis of GoDARTS and SHARE.

### MDLD and Cancer Death

The relationship between MDLD and cancer death was analysed in GoDARTS. In a CPH model adjusted for age, sex, diabetes, BMI and smoking, MDLD was associated with increased risk of cancer death. (HR = 1.40(1.21-1.61), p < 0.0001) FLI was associated with increased cancer death risk in the same CPH model.. (HR = 1.009(1.002 - 1.015), p =9.8×10^−3^)

MDLD was associated with increased risk of non-cancer death in the same model. (HR = 1.23(1.12-1.35), p < 0.0001) To estimate the effects of MDLD specifically on cancer death more accurately, competing risks analyses were run.

A CRR using Fine and Grays’s method was run to analyse the association between MDLD and cancer death with non-cancer related death as a competing risk. In a model with sex, T2DM, smoking, obesity and age, MDLD increased risk of cancer with a subdistribution hazards ratio (SHR) of 1.28 (95% CI =1.11 - 1.47, p < 0.0001).

In SHARE, a CRR adjusted for sex and T2DM with non-cancer death as a competing risk was run. Patients with MDLD had a significantly higher risk of cancer death. (SHR = 3.12 (2.38– 4.10), p <0.0001)

In those patients who died during the follow up period of GoDARTS, MDLD was associated with increased chance of cancer being the main cause of death in a logistic regression adjusted for age, sex, T2DM, smoking and BMI (OR = 1.33 (1.10 – 1.62), p = 3.6×10^−3^). This was also found in SHARE in a logistic regression adjusted for sex and T2DM. (OR = 1.54(1.17 – 2.03), p = 2.0×10^−3^)

Further analysis showed that this association between MDLD and cancer death is one of the major drivers of the shorter life expectancies of MDLD patients. This is shown in the supplementary appendix. The proportion of all deaths with cancer as the main or a contributing cause in GoDARTS are shown in Table 4.

**Table 4:** Proportion of All Deaths Due to Cancer Stratified by NAFLD and Type 2 Diabetes Status in GoDARTS. Table 4 shows the proportion of all deaths which are attributable to cancer. For example, in patients with NAFLD and not diabetes, 41.25% of deaths had cancer a the main cause, and 45.26% of all deaths in this group had cancer as a main or contributing cause.

| Group | Cancer as Main Cause of Death Rate | Cancer as Contributing Cause of Death Rate | Total Number in Group |
| --- | --- | --- | --- |
| No NAFLD or T2DM | 31.33% | 35.34% | 382 |
| T2DM | 24.28% | 30.29% | 449 |
| NAFLD | 41.25% | 45.26% | 559 |
| Both NAFLD and T2DM | 27.80% | 31.73% | 1853 |

Further to the analyses in GoDARTS and SHARE, similar results were found in the Tayside and Fife diabetics’ cohort. MDLD was associated with increased cancer death. (SHR = 1.40(1.20-1.63), p < 0.0001) Full results are shown in the supplementary appendix.

### *PNPLA3* and Cancer Incidence

The effects of *PNPLA3* on cancer incidence during the follow up period in GoDARTS and SHARE were evaluated. Homozygous carriers of *PNPLA3* rs738409 had increased risk of cancer incidence (HR = 1.27 (1.02-1.58), p = 3.1×10^−2^). These results were meta analysed with results from SHARE, shown in Figure 3.

**Figure 3:**
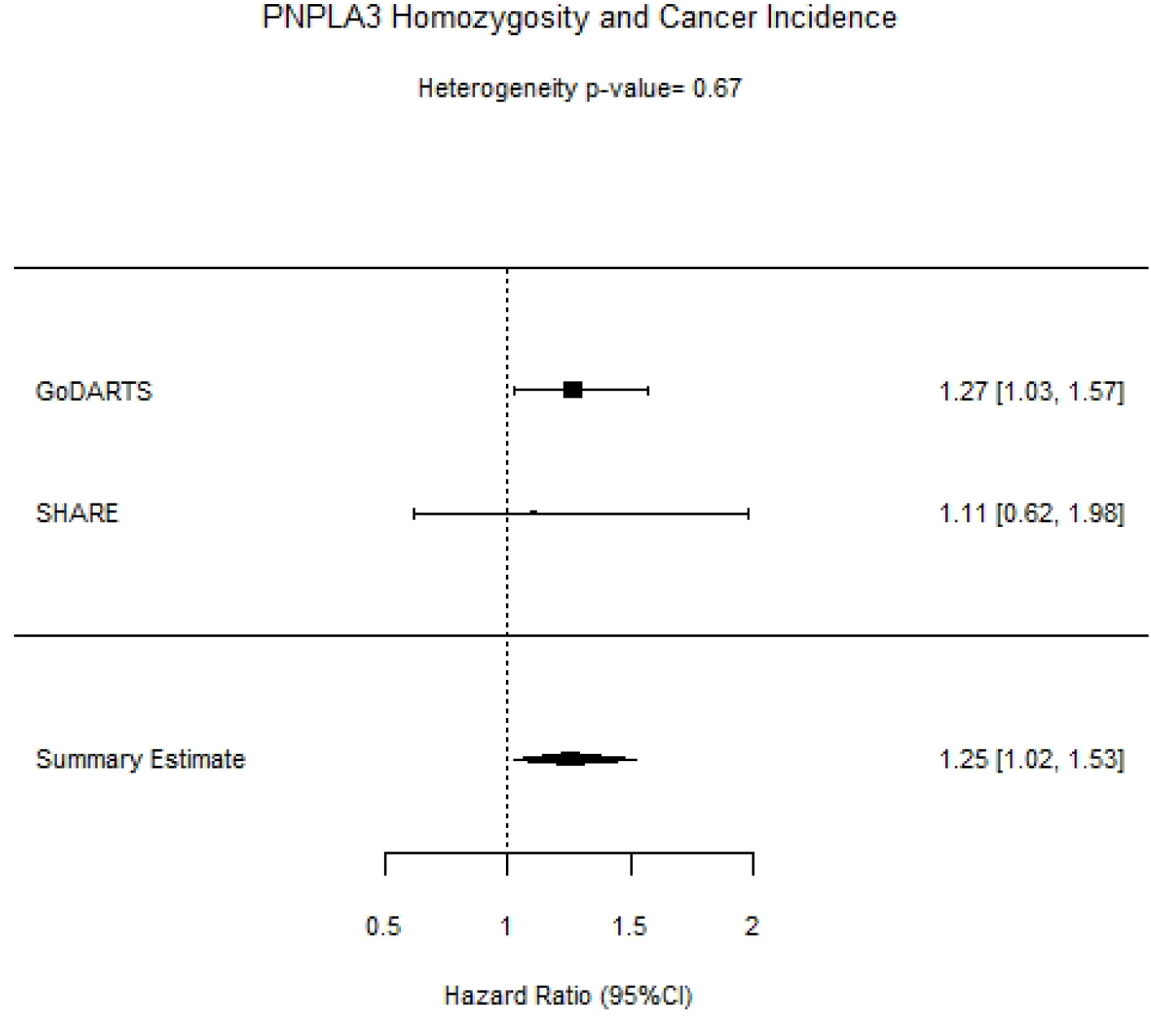
Forest plot of Cancer Incidence and *PNPLA3*rs738409 in GoDARTS, SHARE, and meta-analysis of both cohorts.

This association was also observed in GoDARTS when patients with liver cancer were excluded from analysis, as *PNPLA3* rs738409 has been shown to increase liver cancer risk.^38^ (HR = 1.26(1.01-1.58), p = 3.8×10^−2^) Similar results were found in an adjusted CRR with death as a competing risk. (SHR =1.24(1.00-1.54), p = 4.9×10^−2^)

Mendelian randomisation analysis was conducted to estimate the effect of MDLD on cancer incidence. Using the ratio method in a meta analysis of GoDARTS and SHARE, MDLD was found to be significantly associated with cancer incidence, with a β estimate of 1.33(95% CI = 0.18 - 2.49, p = 0.023)

## Discussion

### Summary of Key Results

In this study, we found that a significant increase in cancer incidence exists in patients with MDLD, high FLI, high FIB-4 and NASH. We also report an association between the strongest genetic instrument for NAFLD risk (*PNPLA3* rs738409) and cancer incidence. Cancer incidence and death was higher in those who had MDLD in GoDARTS, SHARE, and T&F using the raised ALT definition as a surrogate of NAFLD. This demonstrates the generalisability of this result. This is the first truly large scale observational study to show these associations, as well as the first to show the effect of BMI on cancer incidence is driven to null when MDLD is accounted for. In SHARE and T&F, NAFLD admissions were associated with increased cancer incidence. NASH also increased cancer incidence, with a larger effect size than MDLD. Other non-invasive biomarkers including Fatty Liver Index and FIB-4 score prior to enrolment to the GoDARTS study were also found to increase risk of cancer during the follow-up period. These results support findings from other published studies that link NAFLD to cancer of all types. ^3,4^ It also suggests that the more pro-inflammatory form of NAFLD, NASH, may have more of an effect and this may give clues to the biological mechanism(s).

### Limitations

#### MDLD Phenotype

The MDLD phenotype may be a limitation of this study. Case ascertainment for NAFLD is clinically performed using ultrasound, or the gold standard biopsy method. However, these tests are not routinely performed in order to diagnose NAFLD. Therefore, in a population cohort, data on liver biopsies and ultrasound scans are not commonly found. Instead, chronically raised ALTs in the absence of virological, immunoligcal and alcoholic liver insult is generally understood to indicate NAFLD.^14,39^ There is substantial evidence linking ALT levels to NAFLD, but also evidence that NAFLD can exist in patients with normal ALT levels, however the true normal range used in this study negates this.^23^ Furthermore, a non-sensitive MDLD phenotype would drive the association towards null, and therefore we cannot exclude the fact that the true association may be stronger than that we have observed. While we acknowledge that ALT levels may have a limited sensitivity for defining mild NAFLD, we have shown that our ALT based definition is highly sensitive for more advanced cases, such as those with the Fatty Liver Index measured and those hospitalised with steatosis. In GoDARTS, SHARE and T&F, we estimated sensitivity to be 97.4%, 75.3% and 94.6% respectively for such advanced cases. Indeed, our use of exposures such as MDLD, FIB-4, FLI and NASH are included under the umbrella of metabolic associated fatty liver disease (MAFLD). ^16^

Genetic evidence for the suitability of our phenotype ascertainment is demonstrated by the observation that the major NAFLD susceptibility variant in *PNPLA3*, rs738409, was associated with our MDLD phenotype with an to a very similar magnitude to that previously reported. ^40^ The high sensitivity of the phenotype and similar effects of other NAFLD related phenotypes on cancer incidence, plus previous literature linking NAFLD to cancer support the validity of the ALT based MDLD phenotype. ^3,4^

Whilst we show that our MDLD phenotype is accurate, even if part of the aetiology of the raised ALT levels is alcohol or another cause, this is still an important and interesting result. The observation that when ALT levels are taken into account, BMI no longer associates with cancer incidence changes current understanding of the link between cancer and obesity.

We found NASH to be associated with increased cancer incidence, and suggest its associated hepatic inflammation may contribute to cancer risk. The majority of patients with NASH however also have a diagnosis of fibrosis, which could mean the effect is fibrosis rather than inflammation driven.

#### Covariate Data Missingness

The missingness of BMI and smoking data for patients in SHARE is a possible limitation of the current study. In the analysis of cancer incidence in GoDARTS, T&F and the sub-group of SHARE patients with BMI data available, the inclusion of BMI as a covariate did not modify the association between MDLD and cancer. In GoDARTS also, MDLD was not associated with rates of smoking when age and sex were adjusted for. Due to this, the analysis of MDLD and cancer without BMI and smoking as covariates is still valid, and comparable with the analyses undertaken in GoDARTS. Allen et al, used similar methodology, as they did not correct for smoking and found that BMI played a relatively small part in cancer risk compared to NAFLD.^4^ The self-reported nature of alcohol intake in GoDARTS, and missingness of this data in SHARE and T&F, as well as the ubiquitous nature of alcohol consumption at the sub clinical level, does not allow us to exclude the possibility that general alcohol consumption may play a role in the relationship between NAFLD and cancer, however this is likely to be a limitation of the concept of NAFLD in general.

#### BMI as a marker of Obesity

BMI is an accurate and useful marker of obesity, although is not perfectly correlated with abnormal body fatness as factors such as muscle mass can impact the result.^41^ To assess whether this was a factor in the lack of association between BMI and cancer incidence, other measures of body fatness including waist measurement and weight were analysed. These also did not associate with cancer incidence when MDLD was taken into account.

### Interpretation of Results

Hepatocellular carcinoma has long been associated with NAFLD and is widely recognised as one of the most severe endpoints of NAFLD.^2^ There is emerging evidence that the association between NAFLD and cancer extends beyond the liver to other parts of the body. Kim et al. found in a cohort follow-up study that, in addition to an increased risk of liver cancer, NAFLD dramatically increased rates of extrahepatic cancers, including breast and colon in those who were diagnosed with NAFLD prior to the 10 year follow-up period.^3^ Allen et al similarly showed that NAFLD was associated with increased extrahepatic cancer risk, in sites such as the colon, lung and prostate.^4^ In the current study, we found an increase in cancer incidence in many of these specific sites, including breast, colon, liver, lung and prostate. Collectively, these data, including the results that we describe, supports the notion that NAFLD increases incident cancer risk.

We found that MDLD was also associated with increased risk of cancer death in all three cohorts. This data correlates with our earlier findings that NAFLD is associated with increased cancer incidence, as increased incidence is naturally linked to increased mortality. Analysis of causes of death as reported by ICD10 codes in medical records showed that the deaths of patients with MDLD were more likely to be due to cancer. Cancer was also responsible for a large amount of the shorter lifespans of patients with MDLD, as there was no significant effect of MDLD on age of death when patients with a cancer diagnosis were excluded. Similar results were found in a recent study in a large Swedish cohort with biopsy confirmed NAFLD.^42^ In this study, Simon et al. found that excess death in NAFLD patients was primarily driven by extra-hepatic cancers and cirrhosis, while other causes such as cardiovascular disease and HCC had only a small effect. These findings agree with those of the current study, further implicating NAFLD in the development of extrahepatic cancer.

We showed that homozygous carriers of the *PNPLA3* NAFLD risk variant, rs738409, had an increased risk of cancer incidence. In a Mendelian randomisation analysis, we showed *PNPLA3* rs738409 increased MDLD incidence, MDLD increased cancer incidence, and *PNPLA3* rs738409 increased cancer incidence. This novel finding is supporting evidence that MDLD is causally associated with increased cancer incidence.

Substantial evidence links cancer to hyperinsulinemia. For example, hyperinsulinemia has been found to be a risk factor for colon cancer.^43^ Patients with NAFLD are more likely to have hyperinsulinemia, and this is associated with reduced insulin clearance.^44^ This insulin excess may underlie, at least in part, the mechanistic basis by which NAFLD increases cancer incidence, as insulin/Igf-1 may promote tumour formation through mitogenic pathways downstream of their receptors.^45^ Furthermore, NAFLD is a pro-inflammatory state that may inhibit cell cycle checkpoints. The larger effect sizes of NASH and FIB-4 on cancer incidence observed in this study are consistent with the notion of inflammation driving a proportion of cancer risk. Indeed, this observation has been validated recently in a study by Pfister et al., which showed inhibited anti-tumour surveillance in those with HCC caused by NASH.^11^

In a model adjusted for age and sex, BMI was found to be associated with increased cancer incidence. Many studies have shown increased cancer risk with increasing BMI, therefore this finding is consistent with previous literature. We found that BMI was not associated with overall cancer incidence when NAFLD was taken into account, and the same was found for waist and weight measurements. We also found that individuals who were obese but did not have MDLD were not at increased risk of cancer incidence compared to those of a healthy weight. This finding supports those of Allen et al.^4^ When we limited analyses to so-called obesity related cancers, we found similar results, as BMI was associated with cancer incidence, but not when MDLD was adjusted for. This was found in all three cohorts analysed. The lack of independent association between BMI and cancer incidence in our study may suggest that NAFLD is a major component in the increased risk of cancer observed in overweight and obese patients.

### Generalisability

The results of this study are generalisable to those of white European descent, and especially those who are at increased risk of NAFLD including patients with obesity and T2DM. Given the high prevalence of obesity, T2DM and other metabolic dysfunctions, the results have important implications for a large portion of the population. The large number of participants involved allowed for high statistical power further ensuring the external validity of results. The use of population level data in the T&F replication limited the influence of selection bias on results.

## Conclusion

In the current study we have shown that MDLD is associated with increased risk of cancer incidence. There is also an association between MDLD and cancer death, and cancer is a key factor in the shorter life expectancies associated with MDLD patients. Furthermore, we are first to show the association between BMI and cancer is driven to null when MDLD is included in the model. This is further replicated in two additional, large cohorts, demonstrating the robust nature of this relationship. Given the large numbers of participants, these findings are likely generalisable to the general population. A key, novel finding of the study was that the missense variant *PNPLA3* rs738409 is associated with increased cancer incidence. These findings suggests that the effect of NAFLD on cancer incidence may be causative, and that a major component of the association between body weight and cancer may be driven by NAFLD.

## Data Availability

Data analysed in this manuscript is not publicly available.

## Acknowledgements and Ethics

We are grateful to all the participants in this study, the general practitioners, the Scottish School of Primary Care for their help in recruiting the participants and to the whole team, which includes interviewers, computer and laboratory technicians, clerical workers, research scientists, volunteers, managers, receptionists and nurses. The study complies with the Declaration of Helsinki. We acknowledge the support of the Health Informatics Centre, University of Dundee, for managing and supplying the anonymized data and NHS Tayside, the original data owner. The NHS provided ethical approval for access to the anonymised data. Dissemination of the results of this study to participants was not applicable. We acknowledge the support of the NIHR for data acquisition. SHARE has ongoing funding from NHS Research Scotland and established by funding fromThe Wellcome Trust Biomedical Resource GrantNo. 099177/Z/12/Z. The Wellcome Trust United Kingdom Type 2 Diabetes Case Control Collection (GoDARTS) was funded by The Wellcome Trust (072960/Z/03/Z, 084726/Z/08/Z, 084727/Z/08/Z, 085475/Z/08/Z, 085475/B/08/Z) and as part of the EU IMI-SUMMIT program.

## References

1. Younossi Z, Loomba R, Rinella M, Bugianesi E, Marchesini B. Current and Future Therapeutic Regimens for Non-alcoholic Fatty Liver Disease (NAFLD) and Non-alcoholic Steatohepatitis (NASH). Hepatology 2017;(5):1–36.

2. Michelotti GA, Machado M V, Diehl AM. NAFLD, NASH and liver cancer. Nat Rev Gastroenterol & Hepatol [Internet] 2013;10:656. Available from: https://doi.org/10.1038/nrgastro.2013.183

3. Kim GA, Lee HC, Choe J, et al. Association between non-alcoholic fatty liver disease and cancer incidence rate. J Hepatol [Internet] 2018;68(1):140–6. Available from: https://doi.org/10.1016/j.jhep.2017.09.012

4. Allen AM, Hicks SB, Mara KC, Larson JJ, Therneau TM. The Risk of Incident Extrahepatic Cancers is higher in Nonalcoholic Fatty Liver Disease than Obesity - a Longitudinal Cohort Study. J Hepatol 2019;

5. Younossi ZM, Stepanova M, Negro F, et al. Nonalcoholic Fatty Liver Disease in Lean Individuals in the United States. Medicine (Baltimore) [Internet] 2012;91(6):319–27. Available from: http://content.wkhealth.com/linkback/openurl?sid=WKPTLP:landingpage&an=00005792-201211000-00003

6. Gaggini M, Morelli M, Buzzigoli E, DeFronzo RA, Bugianesi E, Gastaldelli A. Non-alcoholic fatty liver disease (NAFLD) and its connection with insulin resistance, dyslipidemia, atherosclerosis and coronary heart disease. Nutrients 2013;5(5):1544–60.

7. Brooke Steele C, Thomas CC, Jane Henley S, et al. Vital signs: Trends in incidence of cancers associated with overweight and obesity — United States, 2005-2014. Morb Mortal Wkly Rep 2017;66(39):1052–8.

8. Wolin KY, Carson K, Colditz GA. Obesity and cancer. Oncologist 2010;15(6):556–65.

9. Secretan BL, Ph D, Scoccianti C, Ph D, Loomis D, Ph D. Spe ci a l R e p or t Body Fatness and Cancer — Viewpoint of the IARC Working Group. 2019;

10. Polyzos SA, Kountouras J, Mantzoros CS. Adipokines in nonalcoholic fatty liver disease. Metabolism 2016;65(8):1062–79.

11. Pfister D, Núñez NG, Pinyol R, et al. NASH limits anti-tumour surveillance in immunotherapy-treated HCC. Nature 2021;592(7854):450–6.

12. Sanyal D, Mukherjee P, Raychaudhuri M, Ghosh S, Mukherjee S, Chowdhury S. Profile of liver enzymes in non-alcoholic fatty liver disease in patients with impaired glucose tolerance and newly detected untreated type 2 diabetes. Indian J Endocrinol Metab 2015;19(5):597–601.

13. Alexander M, Loomis AK, Fairburn-Beech J, et al. Real-world data reveal a diagnostic gap in non-alcoholic fatty liver disease. BMC Med 2018;16(1):1–11.

14. Sattar N, Forrest E, Preiss D. Non-alcoholic fatty liver disease. BMJ [Internet] 2014;349. Available from: https://www.bmj.com/content/349/bmj.g4596

15. Mofrad P, Contos MJ, Haque M, et al. Clinical and histologic spectrum of nonalcoholic fatty liver disease associated with normal ALT values. Hepatology 2003;37(6):1286–92.

16. Eslam M, Sanyal AJ, George J, et al. MAFLD: A Consensus-Driven Proposed Nomenclature for Metabolic Associated Fatty Liver Disease. Gastroenterology [Internet] 2020;Available from: http://www.sciencedirect.com/science/article/pii/S0016508520301712

17. Eslam M, Newsome PN, Sarin SK, et al. Expert Opinion A new de fi nition for metabolic dysfunction-associated fatty liver diseaseLJ: An international expert consensus statement. J Hepatol [Internet] 2020;73(1):202–9. Available from: https://doi.org/10.1016/j.jhep.2020.03.039

18. Wong CA, Araneta MRG, Barrett-Connor E, Alcaraz J, Castañeda D, Macera C. Probable NAFLD, by ALT levels, and diabetes among Filipino-American Women. Diabetes Res Clin Pract 2008;79(1):133–40.

19. Morris AD, Boyle DI, MacAlpine R, et al. The diabetes audit and research in Tayside Scotland (DARTS) study: electronic record linkage to create a diabetes register. DARTS/MEMO Collaboration. BMJ 1997;315(7107):524–8.

20. McKinstry B, Sullivan FM, Vasishta S, et al. Cohort profile: The Scottish Research register SHARE. A register of people interested in research participation linked to NHS data sets. BMJ Open 2017;7(2):1–8.

21. Younossi ZM, Koenig AB, Abdelatif D, Fazel Y, Henry L, Wymer M. Global epidemiology of nonalcoholic fatty liver disease-Meta-analytic assessment of prevalence, incidence, and outcomes. Hepatology 2016;64(1):73–84.

22. WHO | International Classification of Diseases, 11th Revision (ICD-11). WHO 2019;

23. Schindhelm RK, Diamant M, Dekker JM, Tushuizen ME, Teerlink T, Heine RJ. Alanine aminotransferase as a marker of non-alcoholic fatty liver disease in relation to type 2 diabetes mellitus and cardiovascular disease. Diabetes Metab Res Rev 2006;22(6):437–43.

24. Kanth VVR, Sasikala M, Rao PN, Steffie Avanthi U, Rao KR, Nageshwar Reddy D. Pooled genetic analysis in ultrasound measured non-alcoholic fatty liver disease in Indian subjects: A pilot study. World J Hepatol [Internet] 2014;6(6):435–42. Available from: https://pubmed.ncbi.nlm.nih.gov/25018854

25. Verma S, Jensen D, Hart J, Mohanty SR. Predictive value of ALT levels for non-alcoholic steatohepatitis (NASH) and advanced fibrosis in non-alcoholic fatty liver disease (NAFLD). Liver Int 2013;33(9):1398–405.

26. Giannini EG, Testa R, Savarino V. Liver enzyme alteration: a guide for clinicians. C Can Med Assoc J = J l’Association medicale Can 2005;172(3):367–79.

27. Prati D, Taioli E, Zanella A, et al. Updated definitions of healthy ranges for serum alanine aminotransferase levels. Ann Intern Med 2002;137(1):1–10.

28. Ioannou GN, Weiss NS, Boyko EJ, Mozaffarian D, Lee SP. Elevated serum alanine aminotransferase activity and calculated risk of coronary heart disease in the United States. Hepatology 2006;43(5):1145–51.

29. Hickman IJ, Russell AJ, Prins JB, Macdonald GA. Should patients with type 2 diabetes and raised liver enzymes be referred for further evaluation of liver disease? Diabetes Res Clin Pract 2008;80(1):2007–9.

30. Andersson HI, Ejlertsson G, Leden I, Rosenberg C. Chronic pain in a geographically defined general population: studies of differences in age, gender, social class, and pain localization. Clin J Pain [Internet] 1993;9(3):174—182. Available from: http://europepmc.org/abstract/MED/8219517

31. Bedogni G, Bellentani S, Miglioli L, et al. The fatty liver index: A simple and accurate predictor of hepatic steatosis in the general population. BMC Gastroenterol 2006;6:1–7.

32. Sterling RK, Lissen E, Clumeck N, et al. Development of a simple noninvasive index to predict significant fibrosis in patients with HIV/HCV coinfection. Hepatology [Internet] 2006;43(6):1317–25. Available from: https://doi.org/10.1002/hep.21178

33. Marcuccilli M, Chonchol M. NAFLD and chronic kidney disease. Int J Mol Sci 2016;17(4):1–15.

34. Romeo S, Kozlitina J, Xing C, et al. Genetic variation in PNPLA3 confers susceptibility to nonalcoholic fatty liver disease. Nat Genet 2008;40(12):1461–5.

35. Lawlor DA, Harbord RM, Sterne JAC, Timpson N, Davey Smith G. Mendelian randomization: using genes as instruments for making causal inferences in epidemiology. Stat Med 2008;27(8):1133–63.

36. Weisell RC. Body mass index as an indicator of obesity. 2002;11.

37. Fleming M, Kirby B, Penny KI. Record linkage in Scotland and its applications to health research. J Clin Nurs 2012;21(19–20):2711–21.

38. Liu YL, Patman GL, Leathart JBS, et al. Carriage of the PNPLA3 rs738409 C >g polymorphism confers an increased risk of non-alcoholic fatty liver disease associated hepatocellular carcinoma. J Hepatol [Internet] 2014;61(1):75–81. Available from: http://dx.doi.org/10.1016/j.jhep.2014.02.030

39. Sattar N, McConnachie A, Ford I, et al. Serial Metabolic Measurements and Conversion to Type 2 Diabetes in the West of Scotland Coronary Prevention Study. Diabetes [Internet] 2007;56(4):984 LP – 991. Available from: http://diabetes.diabetesjournals.org/content/56/4/984.abstract

40. Wang X, Liu Z, Wang K, et al. Additive Effects of the Risk Alleles of PNPLA3 and TM6SF2 on Non-alcoholic Fatty Liver Disease (NAFLD) in a Chinese Population [Internet]. Front. Genet.. 2016;7:140. Available from: https://www.frontiersin.org/article/10.3389/fgene.2016.00140

41. Adab P, Pallan M, Whincup PH. Is BMI the best measure of obesityLJ? 2018;1274(March):1–2. Available from: http://dx.doi.org/doi:10.1136/bmj.k1274

42. Simon TG, Roelstraete B, Khalili H, Hagström H, Ludvigsson JF. Mortality in biopsy-confirmed nonalcoholic fatty liver disease: results from a nationwide cohort. Gut [Internet] 2020;gutjnl-2020-322786. Available from: http://gut.bmj.com/content/early/2020/10/18/gutjnl-2020-322786.abstract

43. Giovannucci E. Metabolic syndrome, hyperinsulinemia, and colon cancer: A review. Am J Clin Nutr 2007;86(3):836–42.

44. Kotronen A, Westerbacka J, Bergholm R, Pietiläinen KH, Yki-Järvinen H. Liver fat in the metabolic syndrome. J Clin Endocrinol Metab 2007;92(9):3490–7.

45. Draznin B. Mechanism of the mitogenic influence of hyperinsulinemia. Diabetol Metab Syndr 2011;3(1):2–4.

