## Supplementary Materials for "Metabolic Dysfunction Related Liver Disease as a Risk Factor for Cancer"

**Table of Contents**

Supplementary Tables	2

Tayside and Fife Diabetic Cohort Analysis	5

MDLD and Cancer Death in GoDARTS Supplementary Analysis	5

Cohort Descriptions	6

Data Missingness Analysis	7

Supplementary Appendix References	8

**Author List**

Alasdair Taylor PhD^1^ , Moneeza K Siddiqui PhD^1^, Philip Ambery MBChB^2^, Javier Armisen PhD^3^, Benjamin G. Challis PhD^5^, Carolina Haefliger MD^4^, Ewan R. Pearson PhD^1^, Alex S.F. Doney PhD^1^, John F. Dillon MD^6^, Colin N.A. Palmer PhD^1^

^1^Population Health and Genomics, University of Dundee, Dundee, UK

^2^Late Stage Cardiovascular Renal and Metabolic Research, AstraZeneca, Gothenburg, Sweden

^3^Early Clinical Development, Early Cardiovascular Renal and Metabolism, BioPharmaceuticals R&D, AstraZeneca, Cambridge, UK

^4^Centre for Genomics Research, Discovery Sciences, AstraZeneca, Cambridge, UK

^5^Translational Science & Experimental Medicine, Research and Early Development, Cardiovascular, Renal and Metabolism, BioPharmaceuticals R&D, AstraZeneca, Cambridge, UK

^6^Molecular and Clinical Medicine, University of Dundee, Dundee, UK

**Supplementary Tables**

**Consort of Participants in GoDARTS Study**

Final Number of Participants = 13,695

Extract ALT Measurements from Electronic Health Records

(n =18,280)

Exclude those without Baseline Measurements

(n =16,229)

Exclude Patients with Positive Virology Results

(n =16,202)

Exclude those with Positive Immunology Results

(n =14,999)

Exclude those with Alcohol Excess or Morbidity

(n =13,824)

Exclude those with missing data

(n =13,695)

**Consort of Patients in SHARE**

Final Number of Participants = 62,438 (26,891 analysed after age of 60)

Exclude those also in GoDARTS

(n = 63,780)

Extract Biochemical Measurements from Electronic Health Records

(n = 73,024)

Exclude Patients with Positive Virology Results

(n = 69,599)

Exclude those with Positive Immunology Results

(n = 69,449)

Exclude those with Alcohol Excess

(n = 66,369)

Exclude those with Alcohol Excess

(n = 62,438)

Patients older than 60 years

(n = 26,891)

**Consort of Patients in Tayside and Fife**

Final Number of Participants = 16,312 (11,141 analysed after age of 60)

Exclude those also in SHARE

(n = 52,545)

Extract Biochemical Measurements from Electronic Health Records

(n = 89,553)

Exclude Patients with Positive Virology or Immunology Results

(n = 87,554)

Exclude those with Alcohol Excess or Morbidity

(n = 66,532)

Exclude those also in GoDARTS

(n = 59,476)

Exclude those without BMI data

(n = 16,312)

Patients older than 60 years

(n = 11,141)

**Cancer Incidence in GoDARTS, SHARE and T&F** **Supplementary Analyses**

The number of cancer cases split into two groups by MDLD status is shown in Table 1.
Table 1: Number of Cancer Incidents in MDLD and Healthy Patients During Follow-up Period in GoDARTS

|  | Healthy | MDLD |
| --- | --- | --- |
| N | 6,726 | 6,969 |
| All (Total Cancer Incidents) | 2,620 | 3,417 |
| All (Unique Cancer Patients) | 1,244 | 1,550 |
| Lung | 126 | 166 |
| Breast | 93 | 160 |
| Prostate | 174 | 148 |
| Colon | 121 | 163 |
| Skin | 394 | 422 |
| Liver | 20 | 52 |
| Stomach | 24 | 31 |
| Pancreas | 28 | 36 |
| Ovary | 10 | 38 |

The number of cancer cases in SHARE split into two groups by MDLD status is shown in Table 2.

Table 2: Number of Cancer Incidents in SHARE in MDLD and Healthy Patients During Follow-up Period in SHARE.

|  | Healthy | MDLD |
| --- | --- | --- |
| N | 19,035 | 7,856 |
| All (Total Cancer Incidents) | 7,456 | 1,689 |
| All (Unique Cancer Patients) | 4,586 | 1,142 |
| Lung | 279 | 95 |
| Breast | 856 | 308 |
| Prostate | 575 | 98 |
| Colon | 451 | 92 |
| Skin | 2124 | 329 |
| Liver | 30 | 6 |
| Stomach | 38 | 7 |
| Pancreas | 41 | 15 |
| Ovary | 56 | 15 |

The number of cancer cases in T&F split into two groups by MDLD status is shown in Table 3.

Table 3: Number of Cancer Incidents in T&F in MDLD and Healthy Patients During Follow-up Period in SHARE.

|  | Healthy | MDLD |
| --- | --- | --- |
| N | 5,102 | 6039 |
| All (Total Cancer Incidents) | 2,206 | 1,468 |
| All (Unique Cancer Patients) | 1,109 | 710 |
| Lung | 221 | 107 |
| Breast | 174 | 98 |
| Prostate | 71 | 40 |
| Colon | 122 | 59 |
| Skin | 114 | 55 |
| Liver | 41 | 39 |
| Stomach | 33 | 16 |
| Pancreas | 53 | 47 |
| Ovary | 25 | 19 |

**Tayside and Fife Analysis**

Further to the analyses in GoDARTS and SHARE, similar results were found in the Tayside and Fife diabetics’ cohort. These were analysed separately and not included in the meta analysis as this cohort is predominantly diabetic and may have introduced bias to the other more mixed cohorts.

Out of the 11,141 patients analysed, 1,819 had cancer incidents in the follow up period after the age of 60. MDLD was associated with cancer incidence in the follow up period.( HR = 1.16(1.04-1.29), p =5.9x10^-3^) This was analysed a cox proportional hazards model adjusted for sex, BMI, smoking status, and type T2DM. NAFLD hospitalisations were significantly associated with cancer incidence in the same model. (2.04(1.12-3.71), p = 1.9x10^-2^) When analysis was limited to obesity related cancers, BMI did not show any significant association with cancer incidence when MDLD was adjusted for.

Similarly to GoDARTS and SHARE, in a competing risks regression with non-cancer death as a competing risk, adjusted for sex, T2DM, obesity and smoking, MDLD was associated with increased cancer death. (SHR = 1.40(1.20-1.63), p < 0.001)

Patients with MDLD were more likely to die with cancer as the main cause in T&F in a logistic regression adjusted for age, sex, T2DM, smoking and BMI. (OR = 1.44(1.32 – 1.58), p < 0.001)

**MDLD and Cancer Death in GoDARTS Supplementary Analysis**

In GoDARTS, when stratified by cancer death and non-cancer death, MDLD had no effect on age of death in the non-cancer group. MDLD associated with lower death age in those patients who died with cancer as a main cause. (p < 0.001, β = -2.91, 95% CI= (-2.18, -3.63), adjusted R2 = 0.05) MDLD did not have an effect on age of death in those who never had a cancer diagnosis, but associated with lower age of death in those who had a cancer diagnosis at some point. (β = -2.07, 95% CI= (-1.54, -2.60), adjusted R^2^ = 0.07, p < 0.001)

The mean age of death in those who died with cancer as their main cause of death, and those who didn’t stratified by MDLD and T2DM is shown in table 3.

Table 3: Mean Death age versus Cancer Death, MDLD and Type 2 Diabetes. (**●** indicates condition is present*)*

| Cancer as Main Cause of Death | MDLD | T2DM | Mean Death Age | N |
| --- | --- | --- | --- | --- |
|  |  |  | 82.7 | 263 |
| **●** |  |  | 79.2 | 119 |
|  | **●** |  | 83.0 | 328 |
| **●** | **●** |  | 75.9 | 231 |
|  |  | **●** | 80.8 | 340 |
| **●** |  | **●** | 79.9 | 109 |
|  | **●** | **●** | 79.4 | 1340 |
| **●** | **●** | **●** | 76.6 | 513 |

**Cohorts**

Data from the GoDARTS, SHARE and T&F cohorts were used in the current study. These are described below.

*GoDARTS –* *Genetics of Diabetes Audit and Research in Tayside Scotland* ^1,2^

The GoDARTS study was set up to investigate the genetics of T2DM. It uses EMRs directly from the NHS, giving access to admissions, biochemistry, prescribing, death and haematology records to name a few. Through record EMR linkage, data is available for patients as early as 1987. Records are updated automatically, and many years of data are available for most patients.

The cohort is comprised of 18,306 patients, 10,149 of whom have T2DM and 8,157 of whom were healthy controls. The cohort started recruiting in 1998, and the most recent recruitments were made in 2015. Upon sign-up to the GoDARTS study, controls and cases had a number of phenotypic, lifestyle and demographic variables measured. These include BMI, HbA1c, triglycerides, Scottish Index of Multiple Deprivation Score (SIMD10), smoking status and activity level. In the current study, BMI, age, diabetes status, sex and smoking status data were all taken from the GoDARTS baseline file.

Patients’ medical records were used to define phenotypes for MDLD and for cancer. To define MDLD, the biochemistry file was probed for cases of two raised ALT measurements prior to a patients sign up date to GoDARTS. The biochemistry file includes all biochemical measurements from both hospital and GP visits since 1987 for participants of GoDARTS. Data from virology, immunology and admissions were used to exclude patients with other causes of liver disease.

Cancer incident data was found in the cancer register, which is part of the Scottish Morbidity Register.^3^ Admissions files and deaths files were also probed for incidents of cancer.

Data for deaths and the associated cause of death are taken from the deaths file, which is from the National Records of Scotland record of all deaths in Scotland. ^4^

*SHARE - Scottish Health Research Register* ^5^

The SHARE study was set up as a research register in Scotland to aid research in a number of ways. Like the GoDARTS cohort, SHARE relies on participants voluntarily allowing access to their data held by the NHS for research purposes. Unlike GoDARTS, the purpose of the study is not primarily T2DM, and anyone in Scotland over the age of 16 is eligible.

The same data files from the NHS are available as for GoDARTS, thus the phenotype definitions use the same methods and remain consistent. Data for patients in Tayside and Fife was available for the current study, 73,024 patients in total. All biochemical samples such as ALT and GGT were analysed in the same laboratory, so consistency and comparability of results between GoDARTS and SHARE are not affected by inter-laboratory variance.

Unlike GoDARTS, patients did not have phenotypic, lifestyle and demographic variables measured at baseline. This meant that a baseline point between exposure and follow up had to be selected. This meant that BMI and smoking data were not available for the majority of patients.

*Tayside and Fife Diabetics cohort*

This cohort is made up of every individual within Tayside and Fife health board with a diagnosis of T2DM at any point in their life. This uses the same NHS EHRs as GoDARTS and SHARE, therefore a large quantity of longitudinal data is available for each patient. The key strength of this group is that it is population level data and analysis will not suffer from any selection bias. All biochemical samples were analysed in the same laboratory as SHARE and GoDARTS.

From a population of 89,553 patients with T2DM in Tayside and Fife, 52,545 were eligible once exclusions for alternative causes of liver disease and overlap with GoDARTS and SHARE populations were made.

Similarly to SHARE, patients were not phenotyped at a specific sign up point, and all phenotypes had to be constructed from EHRs. The age of 60 was chosen as baseline to mirror the analysis in SHARE.

Patients in this cohort all developed T2DM at some point in their life, with some having T2DM prior to the beginning of available medical records in 1987, and some developing T2DM later, even within the follow up period after the age of 60 years.

**Missingness**

*Missingness in GoDARTS*

In GoDARTS, there was a small amount of missingness in data for BMI, sex, and age.

There were 40 patients with missing BMI, 81 with missing sex data, and 81 with missing age data. Eight patients did not have information available to allow calculation of their age at sign up. This left 13,695 patients suitable for analysis.

*Missingness in SHARE*

Of the variables of interest, there was missing data in the following variables: Smoking and BMI. Patients with missing data for variables were excluded from analysis in which it was required. Patients with BMI and smoking missing were excluded from some sub-analysis in SHARE. 12,150 patients had smoking data available and 1912 patients had BMI data.

*Missingness in Tayside and Fife Diabetics cohort*

After exclusions had been made for patients with alternative liver insults and overlap with GoDARTS and SHARE, there were 52,545 patients eligible. Of these, 16,312 had BMI data available

**Supplementary Appendix References**

1.	Hébert HL, Shepherd B, Milburn K, et al. Cohort profile: Genetics of Diabetes Audit and Research in Tayside Scotland (GoDARTS). Int J Epidemiol 2018;47(2):380-381j.

2.	Morris AD, Boyle DI, MacAlpine R, et al. The diabetes audit and research in Tayside Scotland (DARTS) study: electronic record linkage to create a diabetes register. DARTS/MEMO Collaboration. BMJ 1997;315(7107):524–8.

3.	Fleming M, Kirby B, Penny KI. Record linkage in Scotland and its applications to health research. J Clin Nurs 2012;21(19–20):2711–21.

4.	Quality of National Records of Scotland (NRS) Data on Deaths | National Records of Scotland [Internet]. [cited 2019 Oct 24];Available from: https://www.nrscotland.gov.uk/statistics-and-data/statistics/statistics-by-theme/vital-events/deaths/deaths-background-information/quality-of-nrs-data-on-deaths

5.	McKinstry B, Sullivan FM, Vasishta S, et al. Cohort profile: The Scottish Research register SHARE. A register of people interested in research participation linked to NHS data sets. BMJ Open 2017;7(2):1–8.
